# A Blood-Based Transcriptomic Signature for PTSD Classification Using Machine Learning

**DOI:** 10.64898/2026.05.10.26352854

**Authors:** Shawheen Alipour, Rishika Pamanji, Emina Jamil, Suneetha Yeguvapalli, Kumaraswamy Naidu Chitrala

## Abstract

Post-traumatic stress disorder (PTSD) remains a significant psychiatric burden; despite growing biomarker research, no blood-based molecular diagnostic tool has been clinically validated for routine use. In this study, we developed a machine learning classifier for PTSD using peripheral blood leukocyte RNA-seq data from combat-exposed U.S. Marines (GSE64813), diagnosed via the Clinician-Administered PTSD Scale (CAPS) under DSM-IV criteria. Differentially expressed genes (DEGs) were identified and further refined through additional filtering criteria, yielding a 90-gene feature set used to train and compare multiple machine learning models. The support vector machine (SVM) classifier achieved the best performance, with an accuracy of 89% and an AUC of 0.95, outperforming logistic regression and random forest approaches. Furthermore we evaluated our model on independent external datasets to assess generalizability. These findings highlight the promise of transcriptomic signatures as a foundation for objective, blood-based PTSD diagnostics, while emphasizing the critical need for robust cross-dataset generalizability.

**Code availability:** https://www.kaggle.com/code/persianexxx/ptsd-final

## Introduction

PTSD is a debilitating psychiatric condition affecting millions of individuals worldwide following exposure to traumatic events. Large-scale epidemiological surveys have estimated its lifetime prevalence at approximately 3.9% globally, with substantially higher rates among conflict-exposed populations [1]. Among war survivors, the burden is particularly severe: a recent meta-analysis estimated that more than 300 million individuals in war-affected regions suffer from PTSD or major depressive disorder [2]. Despite this scale, diagnosis remains entirely clinical, relying on structured interviews such as the Clinician-Administered PTSD Scale (CAPS), which are resource-intensive and subject to reporting bias. The development of objective biological markers capable of identifying at-risk individuals or confirming diagnosis would represent a significant clinical advance.

Peripheral blood has emerged as a promising biomarker source, as it is minimally invasive and reflects the systemic immune and neuroendocrine dysregulation associated with PTSD [3]. Early transcriptomic studies in combat-deployed Marines demonstrated that pre-deployment gene expression profiles could predict subsequent PTSD risk [4], and that post-deployment RNA-seq signatures from peripheral blood leukocytes could discriminate PTSD cases from resilient controls [5]. Breen et al. [6] extended these findings using weighted gene co-expression network analysis (WGCNA), identifying dysregulated innate immune and interferon signalling modules both pre- and post-deployment, replicated across two independent Marine cohorts. Subsequent multi-cohort analyses confirmed consistent enrichment of toll-like receptor and innate immune pathways across veteran populations [7], and transcriptome-wide RNA-seq of World Trade Center responders similarly implicated inflammatory pathways in a civilian PTSD context [8]. Most recently, Daskalakis et al. [9] conducted a landmark multiomics study across brain regions and blood, confirming sex-specific molecular signatures and a central role for interferon signalling in PTSD pathophysiology. Collectively, these studies establish dysregulated innate immune responses in peripheral blood as a reproducible and biologically meaningful feature of PTSD.

The integration of machine learning with high-dimensional transcriptomic data offers a principled framework for exploiting these signatures diagnostically. Schultebraucks and Galatzer-Levy [10] highlight that robust ML workflows require supervised training, cross-validation, and independent dataset validation to yield reliable inference from complex biological data. In practice, SVMs and ensemble methods have demonstrated consistently strong performance across psychiatric transcriptomic classification tasks [11, 12]. However, most existing models are trained and evaluated within a single cohort, and classification accuracy can degrade substantially when transferred across datasets [13], leaving cross-dataset generalizability largely unaddressed. Yu et al. [14] developed an SVM classifier for major depressive disorder from peripheral blood transcripts, achieving 90.6% cross-validated sensitivity and specificity — a directly analogous approach to the present study. Zhu et al. [15] similarly applied computational modelling to PTSD blood transcriptomes, identifying ferroptosis-related genes as candidate biomarkers. Systematic differences in sequencing platforms, library preparation, and normalization pipelines introduce batch effects that can dominate biological signal when classifiers are transferred across cohorts, and standard correction methods such as ComBat [16], while widely used, do not fully resolve these distributional differences.

In this study, we leverage the GSE64813 RNA-seq dataset from combat-exposed U.S. Marines to build a PTSD classifier from differentially expressed genes, comparing Random Forest, Logistic Regression, and SVM under stratified cross-validation. The best-performing model is evaluated on an independent external cohort (GSE97356) [17], with explicit batch correction applied to assess cross-dataset portability. Our results demonstrate the potential of transcriptomic signatures for objective PTSD stratification while highlighting the various challenges that must be addressed for cross-cohort deployment.

## Methods

### Data

Gene expression data were drawn from the Marine Resiliency Study II (MRS II) [6]. The dataset consists of whole-blood RNA-Seq gene expression profiles from peripheral blood leukocytes of 188 U.S. male Marines, collected at two time points: one month prior to deployment and three months following deployment to conflict zones in Iraq or Afghanistan. Following quality control and preprocessing, the analytic sample comprised 94 matched pairs (47 PTSD cases, 47 controls), totaling 188 observations across both time points. All participants were symptom-free at pre-deployment (CAPS ≤25). PTSD diagnosis at post-deployment was assessed using the Clinician Administered PTSD Scale (CAPS)[18], with cases defined by partial or full PTSD criteria per DSM-IV. Raw RNA-Seq data are publicly available through the Gene Expression Omnibus (accession number GSE64814).

### Differential Expression Analysis

Differentially expressed genes (DEGs) were identified using GEO2R, a web-based tool provided by the NCBI Gene Expression Omnibus (GEO), applied to the GSE64813 dataset. GEO2R performs differential expression analysis using the limma package [19], comparing PTSD and control groups across all 188 samples. Genes were ranked by adjusted p-value (Benjamini-Hochberg correction), and a significance threshold of *p*_adj_ *<* 0.05 was applied to obtain the final feature set of 90 genes used for downstream classification. The resulting DEGs were predominantly interferon-stimulated genes, consistent with previously reported innate immune signatures in PTSD [6]. Figure 1 shows the mean difference plot generated by GEO2R, illustrating the magnitude and direction of expression changes between PTSD and control groups.

**Figure 1.**
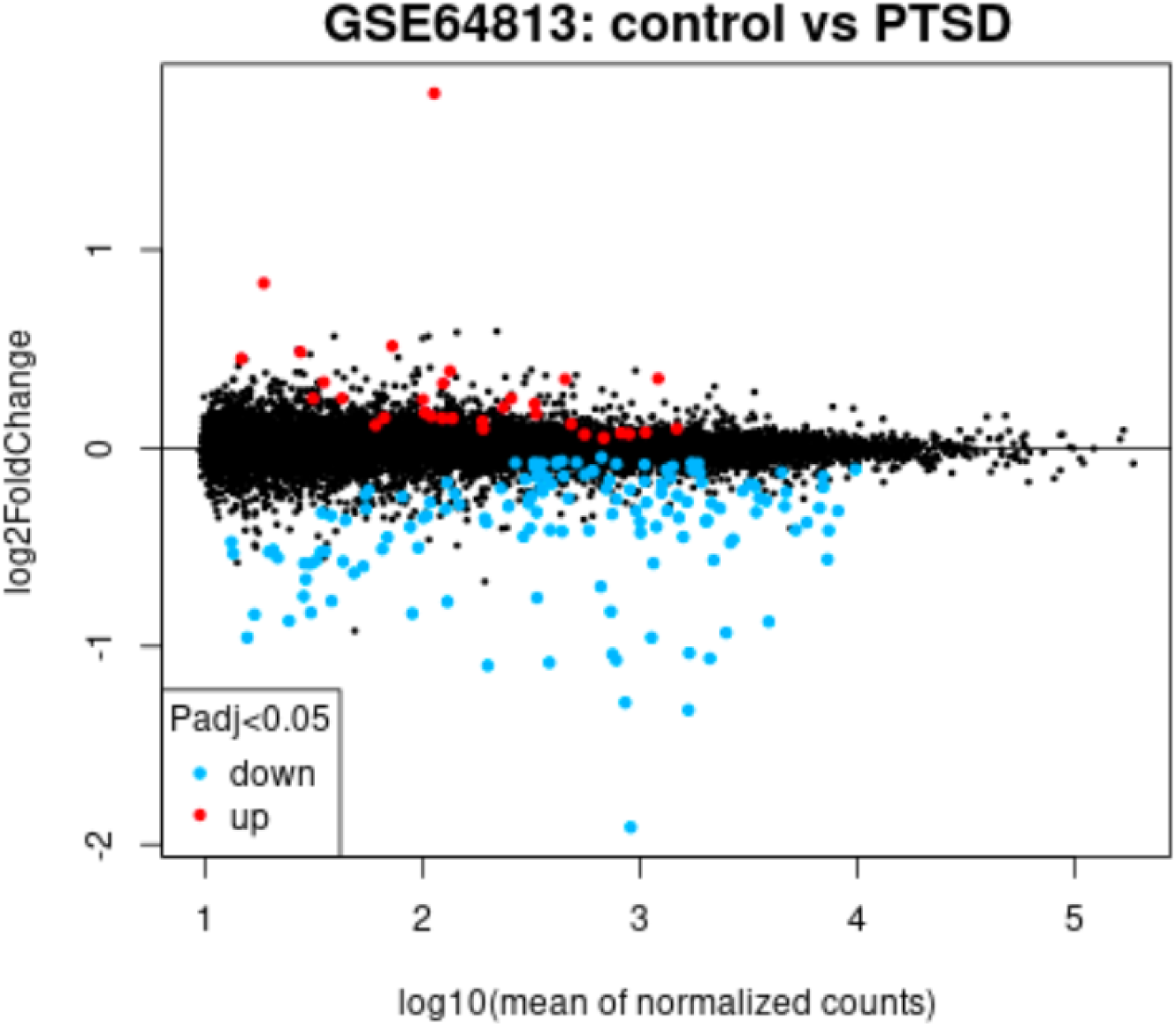
Mean difference plot of differentially expressed genes between control and PTSD groups (GSE64813, *p*_adj_ *<* 0.05), generated via GEO2R.

### Feature Selection

DEGs were filtered from the GEO2R output using a nominal p-value threshold of *p <* 0.05 combined with a minimum absolute log2 fold change of 0.3, yielding a final feature set of 90 genes. These genes were subsequently intersected with those present in the expression matrix, and only the overlapping genes were retained for downstream analysis.

### Machine Learning Classification

The expression matrix was subset to the 90-gene feature set and standardized using `StandardScaler` prior to model training. Three classifiers were evaluated: Random Forest (500 estimators), Logistic Regression (maximum 5000 iterations), and Support Vector Machine (SVM). Model performance was estimated using stratified 5-fold cross-validation (`StratifiedKFold, random state=42`) with accuracy as the scoring metric.

Hyperparameter optimization for the SVM was performed using an exhaustive grid search (`GridSearchCV`) over the following parameter space: regularization parameter *C* ∈{0.1, 1, 10, 100}, kernel ∈{RBF, linear}, and *γ* ∈ {scale, auto} . The optimal configuration was identified as an RBF kernel with *C* = 100 and *γ* = scale, achieving a best cross-validated accuracy of 88.8%. Final model evaluation on the held-out test set was performed using accuracy, precision, recall, F1-score, confusion matrix, and AUC-ROC.

### External Validation

The trained SVM model was externally validated on an independent PTSD dataset (GSE97356). The external dataset was aligned to the 90-gene feature set via column reindexing, with any missing genes imputed using per-gene training set means. To assess and correct for systematic distributional differences between cohorts, kernel density estimation was used to visually compare expression distributions. ComBat batch correction was subsequently applied using the `neuroCombat` package [16], combining training and validation data into a single matrix with batch labels prior to correction. The corrected validation subset was then passed to the trained SVM for final prediction.

## Results

### Differential Expression Analysis

Applying a significance threshold of *p <* 0.05 and a minimum absolute log2 fold change of 0.3 to the GEO2R output yielded predominantly interferon-stimulated genes, including *RSAD2, IFI44L, IFIT1, IFI44*, and *ISG15*, consistent with previously reported innate immune signatures in PTSD [6].

### Model Comparison

Three classifiers were evaluated under stratified 5-fold cross-validation. As shown in Table 1, Logistic Regression achieved the highest mean accuracy among baseline models (78.2%), followed by the linear SVM (77.7%) and Random Forest (71.8%). Following hyperparameter tuning via grid search, the optimized SVM (RBF kernel, *C* = 100, *γ* = scale) achieved a cross-validated accuracy of 88.8%.

**Table 1.** Cross-validated classification performance of evaluated models.

| Model | Mean Accuracy | Std Dev |
| --- | --- | --- |
| Random Forest | 0.718 | 0.088 |
| Logistic Regression | 0.782 | 0.031 |
| SVM (linear kernel) | 0.777 | 0.026 |
| SVM (RBF, tuned)* | 0.888 | 0.030 |

\*Hyperparameters optimized via `GridSearchCV` using the same stratified 5-fold cross-validation scheme.

### SVM Classification Performance

The optimized SVM model was evaluated on a held-out test set of 188 samples (94 PTSD, 94 Control), achieving an overall accuracy of 89%. The classifier demonstrated balanced performance across both classes, with a precision and recall of 0.88 and 0.90 for the control class, and 0.90 and 0.87 for the PTSD class, yielding a macro-averaged F1-score of 0.89. The AUC-ROC was 0.95, indicating strong discriminability between PTSD and control subjects. Figure 2 and Figure 3 show the ROC curve and confusion matrix respectively.

**Figure 2.**
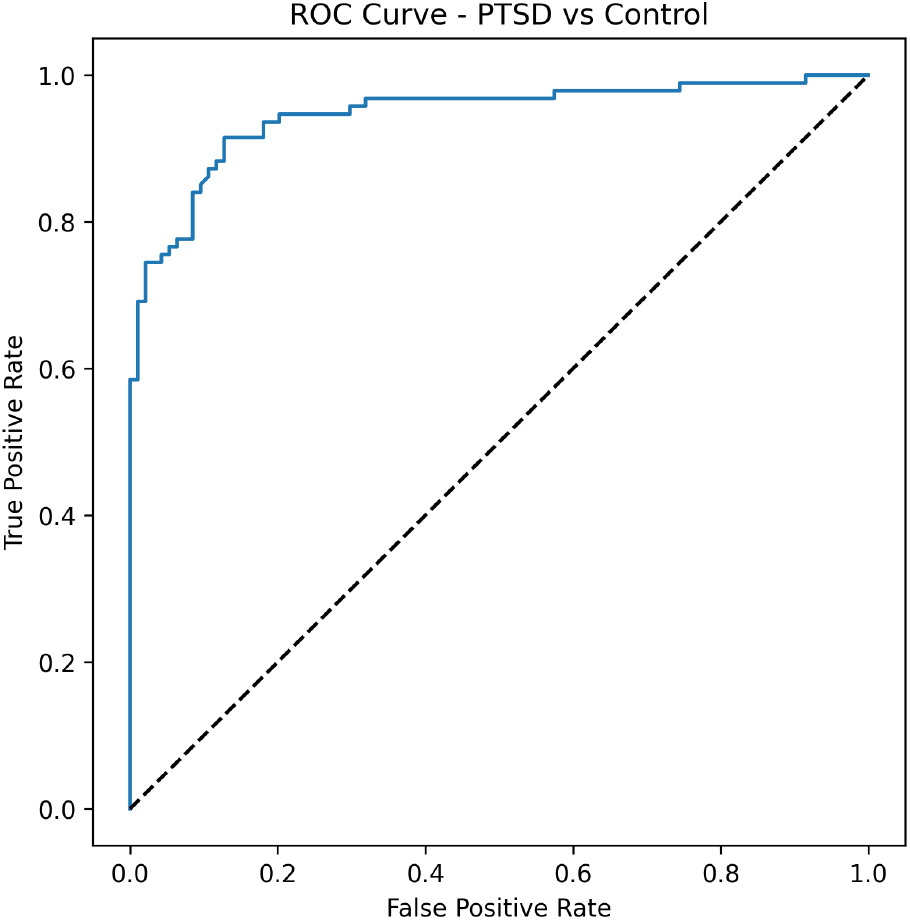
ROC curve of the optimized SVM classifier (AUC = 0.95).

**Figure 3.**
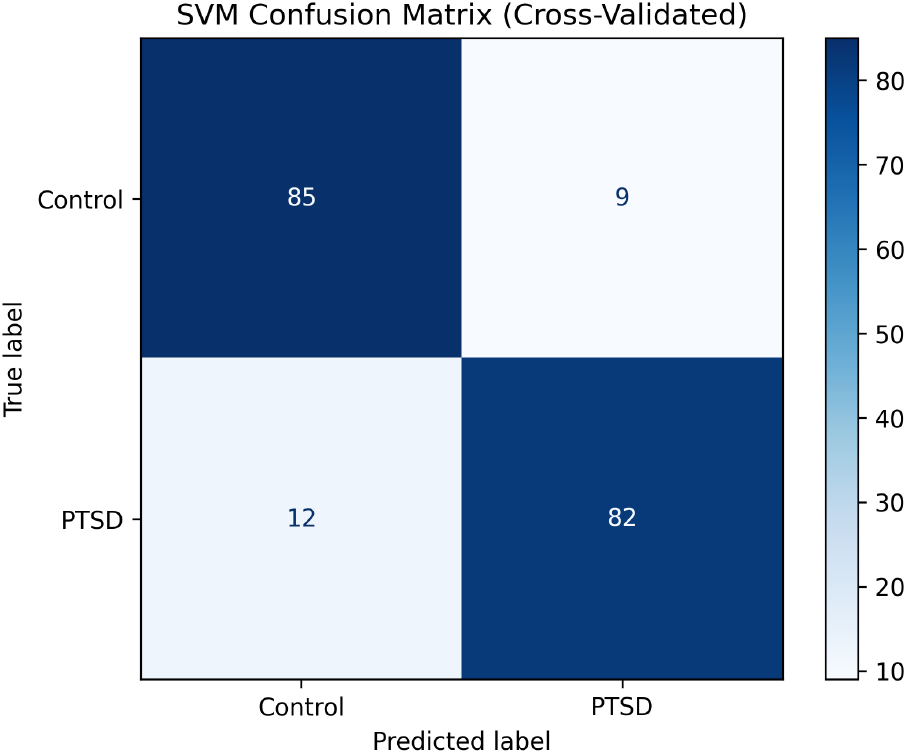
Confusion matrix on the held-out test set (PTSD vs. Control).

### External Validation and Batch Correction

Applying the trained SVM directly to the aligned external dataset (GSE97356) revealed substantial distributional differences between the training and validation cohorts. Figure 4 illustrates the expression divergence across the first 20 samples and 10 genes between the two datasets, highlighting the need for explicit harmonization prior to prediction.

**Figure 4.**
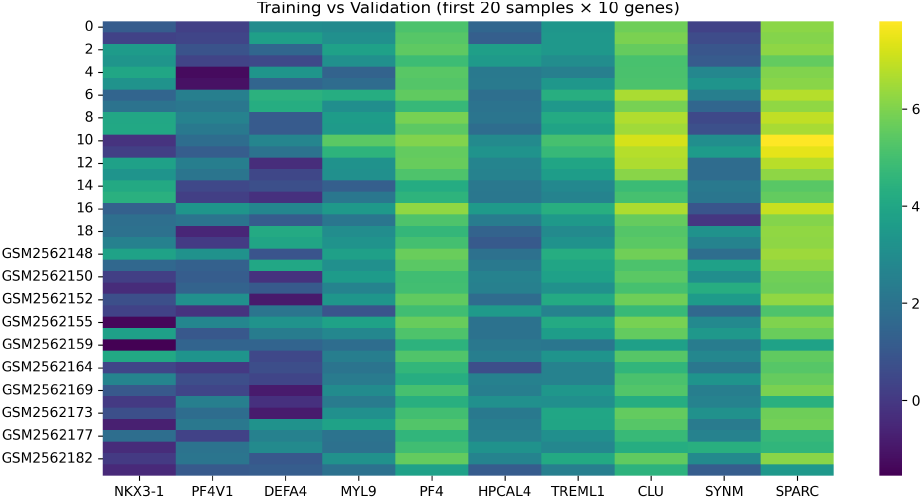
Heatmap comparing expression values of the first 20 samples across 10 genes between training (GSE64813) and validation (GSE97356) cohorts, prior to batch correction.

To address this, three sequential correction strategies were applied and evaluated. First, a global mean shift was applied to the validation set by computing the difference between the training and validation global expression means and adding it as a scalar offset to all validation values. Figure 5 shows the resulting alignment of expression distributions after this correction. Second, quantile normalization was applied to further align the per-gene distributions between cohorts. Finally, ComBat batch correction was applied using the `neuroCombat` package [16], combining training and validation samples into a single matrix with explicit batch labels before correction, then splitting the corrected matrix back into training and validation subsets for prediction.

**Figure 5.**
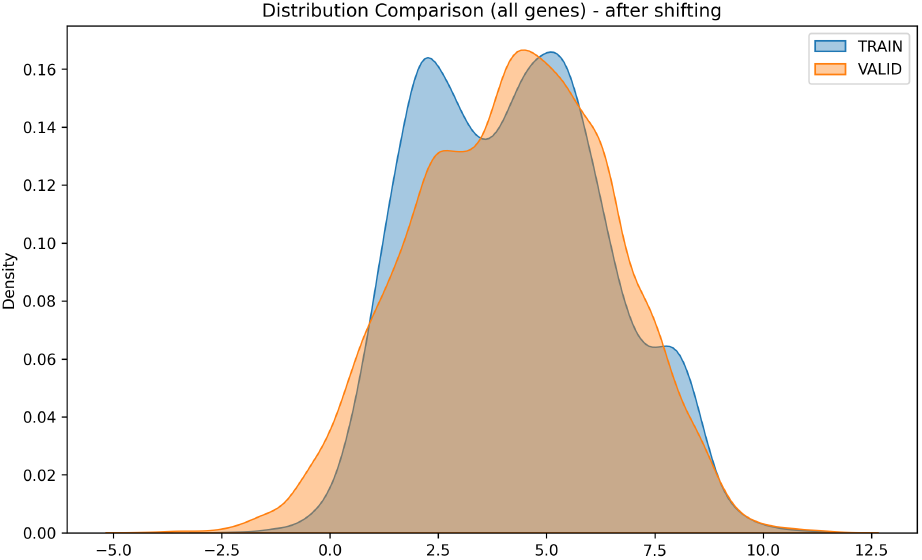
Kernel density estimate of expression distributions for training and validation cohorts after global mean shift correction.

Classification performance degraded substantially relative to cross-validated results, with an overall accuracy of 0.41. The model exhibited a strong bias toward predicting PTSD, achieving a recall of 0.80 for the PTSD class but only 0.17 for controls, reflected in a markedly imbalanced F1-score (PTSD = 0.51, Control = 0.26). These results suggest that the model does not generalise well to external data, likely attributable to differences in expression scale and distribution between datasets, as well as the class imbalance present in the validation set.

## Discussion

Rather than adopting the hub genes identified by Breen et al. directly, an independent DEG analysis was performed to avoid circularity, as those genes were selected on the same dataset. The resulting feature set nonetheless remained biologically consistent with the innate immune and interferon signalling signatures they reported.

Our results demonstrate the feasibility of a blood-based transcriptomic classifier for PTSD, achieving an AUC of 0.95 and 89% accuracy on a held-out test set derived from combat-exposed male U.S. Marines. The dominance of interferon-stimulated and innate immune genes among the top DEGs — including *RSAD2, IFI44L*, and *IFIT1* — is consistent with prior transcriptomic studies in combat-exposed cohorts [4, 5, 6], further reinforcing the role of innate immune dysregulation in PTSD pathophysiology.

The tuned SVM with an RBF kernel and *C* = 100 achieved the strongest classification performance (mean accuracy = 0.888, std = 0.030), outperforming the linear SVM (0.777), logistic regression (0.782), and random forest (0.718). The performance gain over the linear SVM suggests that the relationship between gene expression features and PTSD status is not strictly linearly separable, and that the RBF kernel’s capacity to model non-linear decision boundaries is better suited to this data. Balanced precision and recall across both PTSD and control classes (F1 = 0.89 for both) indicate that the model generalizes evenly and is not biased toward either class, which is particularly important in a clinical context where misclassification of either group carries unnecessary consequences.

Despite promising within-cohort performance, external validation on GSE97356 highlighted the challenge of cross-dataset generalization. Differences in sequencing platforms, normalization pipelines, and sample preparation protocols introduce systematic batch effects that complicate direct model transfer. Although ComBat correction partially mitigated these effects, residual distributional differences between cohorts likely contributed to reduced external performance. This represents a broader and largely unsolved challenge for deploying RNA-seq classifiers across independent datasets, and will be a primary focus of future work.

Notably, these limitations are not unique to transcriptomic approaches — current clinical diagnosis of PTSD relies heavily on self-report instruments and structured interviews, which are vulnerable to stigma-driven underreporting and inter-rater variability, particularly in military populations [20, 21]. Objective, blood-based classifiers offer a complementary diagnostic approach independent of self-report biases [22], motivating continued efforts to overcome the technical barriers that currently limit their cross-dataset generalizability

Several limitations of this study warrant discussion. First, the model was trained and tested exclusively on combat-exposed male U.S. Marines, limiting its generalizability to other populations, including females, civilians, and individuals exposed to non-combat trauma. Second, the relatively small number of publicly available PTSD RNA-seq datasets — many of which differ fundamentally in design, tissue source, or diagnostic criteria — restricts both model training and rigorous external validation. Third, PTSD frequently co-occurs with conditions such as depression, traumatic brain injury, and substance use disorders [23, 24]. The presence of such comorbidities in the cohorts used here is largely uncontrolled for, and may confound the transcriptomic signal attributed to PTSD specifically. Developing classifiers that are robust to psychiatric comorbidities remains an important direction for future work.

Future efforts will focus on several key directions. First, training across multiple independent cohorts using harmonized normalization pipelines will be essential for building classifiers that generalize beyond a single military dataset. Second, expanding validation to more diverse populations — including females, civilians, and survivors of non-combat trauma — is necessary to assess whether the innate immune signature identified here reflects a generalizable biological mechanism or is specific to combat-exposed males. Third, improved cross-platform normalization strategies, beyond ComBat, such as reference gene normalization or deep learning-based domain adaptation, may better address the batch effects that limited external performance here. Finally, incorporating clinical covariates such as comorbid depression, TBI, and substance use into the classification framework could improve both model robustness and biological interpretability, moving toward a clinically deployable transcriptomic tool for early PTSD detection.

## Data Availability

All Data & code available online at:
https://www.kaggle.com/code/persianexxx/ptsd-final

https://www.kaggle.com/code/persianexxx/ptsd-final

